# Patterns and predictors of contraceptive use among post-caesarean women in Sierra Leone: insights from a five-year longitudinal study

**DOI:** 10.64898/2026.03.17.26348588

**Authors:** Maranatha K Banda, Hussein H Twabi, Alex J. van Duinen, Marriott Nliwasa, Michael Kamara, Maria Odland

## Abstract

Caesarean deliveries and short birth intervals predispose to adverse maternal outcomes. Family planning lowers this risk by reducing unplanned pregnancies. This study assessed the uptake and influencing factors of contraceptive use among women at one- and five-years post-caesarean delivery in Sierra Leone.

We performed a secondary analysis of data from a multicentre, prospective observational study involving 1,274 women who underwent caesarean delivery in nine hospitals across Sierra Leone between October 2016 and May 2017. The primary outcome was the use of a modern contraceptive method within five years post-delivery. Multivariable logistic regression analyses were used to identify factors associated with contraceptive uptake.

Overall contraceptive use at five years was 48.5%. The commonest method used at year one was the intrauterine contraceptive device, but this declined significantly from 40.3% to 0.8% by year five (*p* < 0.001). Attending more than two antenatal care visits [aOR 1.96; 95% CI (1.19, 3.23)] and offering a contraceptive method before discharge [aOR 2.44; 95% CI (1.05, 6.40)] were associated with a higher likelihood of modern contraceptive uptake, while delivery at a tertiary/regional facility was associated with a lower likelihood [aOR 0.53; 95% CI (0.34, 0.83)].

Increased contact with the health system was associated with a higher uptake of modern contraceptive methods among post-caesarean women. Strengthening provider–client interaction and integrating contraceptive counselling into routine antenatal and postnatal care could improve contraceptive use and address the unmet need for family planning.

## 1. Background

Globally, the maternal mortality ratio (MMR) has decreased significantly since the early 2000s (1). However, the rate of reduction has slowed down in recent years (2). By 2020, the global MMR remained high at 223 per 100,000 live births (3), raising concerns that the Sustainable Development Goal (SDG) target of fewer than 70 deaths per 100,000 by 2030 may not be met (3).

Sub-Saharan Africa accounts for 70% of global maternal deaths, reflecting inequalities in access to quality healthcare (4). Sierra Leone, located in sub-Saharan Africa, has a maternal mortality ratio of 547 per 100,000 live births (6), more than double the global average. The high mortality ratio underscores the persistent deficiencies in the country’s maternal healthcare delivery. Early childbearing further exacerbates the maternal risk, as one-quarter of adolescents have begun bearing children (5), putting this population at risk of complications such as haemorrhage and eclampsia (6,7).

The mode of birth also influences maternal outcome, and caesarean delivery is associated with more than double the rate of maternal morbidity and mortality compared with vaginal birth (8). Complications from caesarean delivery can occur during the surgery, the postoperative period, or during consecutive pregnancies, such as uterine rupture or placenta accreta (9). A prospective cohort study in Sierra Leone found that 27% of women became pregnant within five years following a caesarean delivery, with those experiencing perinatal loss being four times more likely to conceive again within a year (8). Such patterns contribute to short birth intervals — defined as less than 24 months between births — which are linked to adverse maternal outcomes (10,11). Furthermore, most caesarean deliveries that are done in Sierra Leone are done as emergency surgery and are associated with a high perinatal mortality rate of 190 per 1000 births (12). Therefore, preventing unintended pregnancies is key to reducing maternal deaths (13). Family planning remains a cost-effective intervention that can prevent up to one-third of maternal deaths by reducing unintended pregnancies (13,14).

Under the Free Health Care Initiative, Sierra Leone offers free healthcare, including family planning services, to all pregnant and lactating women (15). The country has committed to increasing its modern contraceptive prevalence rate (mCPR) to at least 32% by 2030 (16). One strategy to achieve this target has been the integration of postpartum family planning (PPFP) into maternal and child services, including antenatal care, delivery, and immunization services (16). Despite these efforts, access to family planning remains hindered by socioeconomic and health system barriers, including frequent stock-outs of medical supplies and a shortage of skilled healthcare workers (17). Moreover, uptake of contraception through the PPFP programme is only 4% (18), indicating that a significant proportion of postpartum mothers are discharged from the health system without receiving a contraceptive method. These challenges are further reflected in the country’s low contraceptive uptake of 29% (18) and a stagnated unmet need for contraception at 25% (7,8).

Given the elevated risks associated with caesarean delivery and the critical role of family planning in reducing maternal mortality, this study aims to assess the uptake and associated factors of modern contraceptive use within five years of delivery among women who underwent a caesarean delivery in Sierra Leone.

## 2. Materials and methods

### Study design

This study utilised secondary data from a prospective five-year observational cohort of women who underwent caesarean delivery in Sierra Leone between 1 October 2016 and 5 May 2017 (12). During the study period, 1,274 women who underwent caesarean delivery with a fetal weight greater than 500 grams across nine study facilities were included in the main cohort. The study sites included four district hospitals in the Northern province (Kabala, Kambia, Magburaka and Port Loko Governmental Hospitals), one regional hospital in the Eastern province (Kenema Governmental Hospital), the national referral centre in the Western province (Princess Christian Maternity Hospital, Freetown), and three private non-profit hospitals (Lion Heart Medical Center and Magbenteh Community Hospital in the Northern province, and Serabu Catholic Hospital in the Southern province).

For this secondary analysis, we included women of all ages who had a caesarean delivery at any of the participating sites. Women were excluded if they underwent a hysterectomy, were lost to follow-up, if a maternal death occurred or if there were errors in data entry (Figure 1).

**Fig 1.**
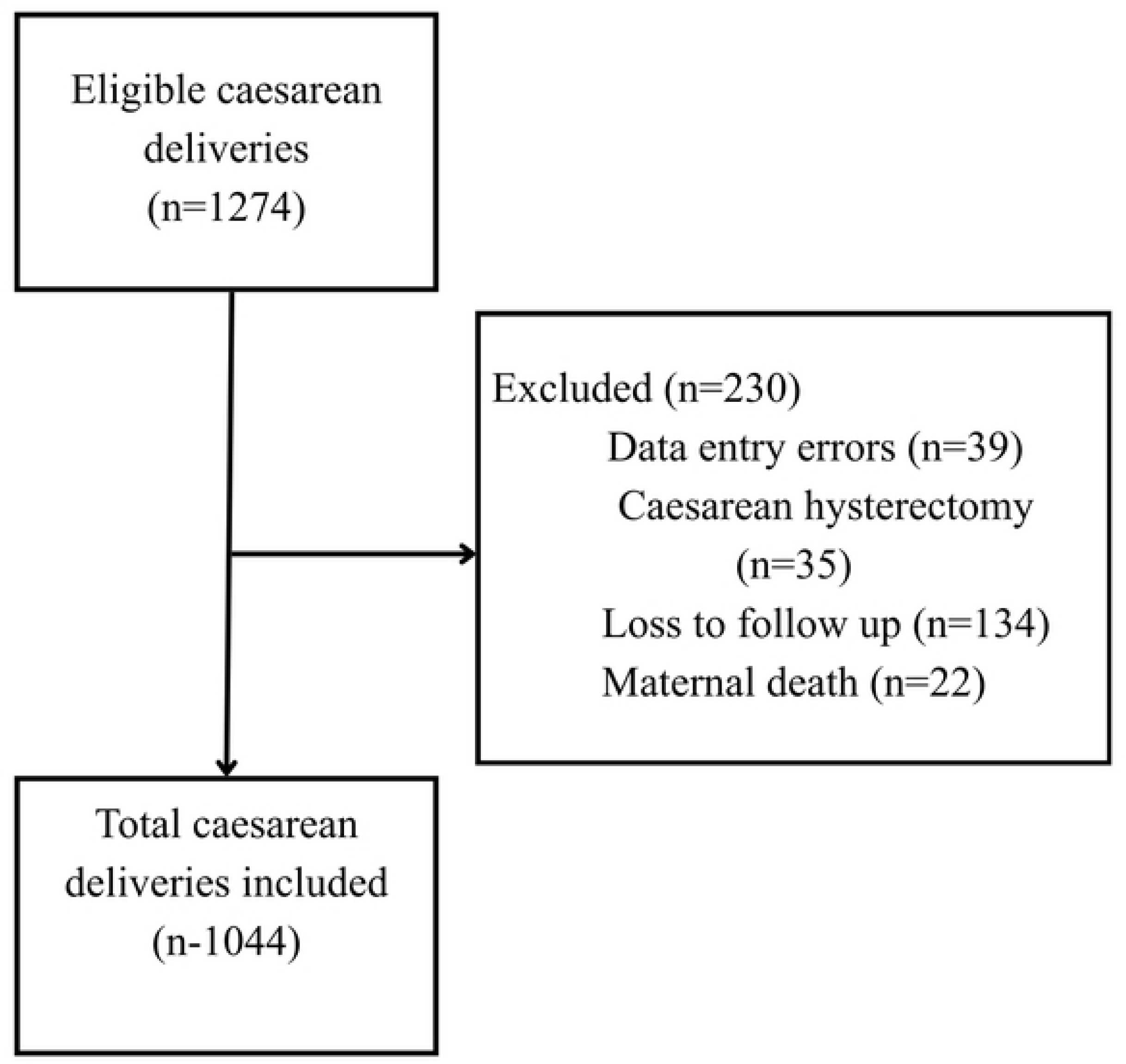
Study flowchart of eligible women post-caesarean delivery in Sierra Leone.

The original study was conducted in accordance with the Declaration of Helsinki and approved by the Sierra Leone Ethics Committee and the Regional Committees for Medical and Health Research Ethics in Central Norway (ethical clearance number 2016/1163). Informed consent was sought before enrolment. For minors, consent was obtained from guardians. Written consent was obtained before or shortly after the caesarean delivery.

### Study variables

#### Dependent variable

The main outcome for this analysis was utilisation of modern contraception following caesarean delivery. This was a binary variable defined by the use of either condoms, depot medroxyprogesterone acetate (DMPA), implant, intrauterine contraceptive device (IUCD), pills, or a bilateral tubal ligation (BTL). The use of DMPA in year one was not captured in the main cohort’s data collection tool. However, this data was captured in year five. For outcome ascertainment, home visits were conducted at 30 days, one year, and five years post-delivery.

#### Independent variables

Nine explanatory variables were selected based on evidence from literature and their availability in the dataset (5,19–21) . The variables included: maternal age, parity, education status, marital status, number of antenatal care (ANC) visits, perinatal death, facility type, wealth quintile and whether a contraceptive method was offered at any point during admission (a composite variable combining pre-operative and pre-discharge offers).

### Data analysis

Complete case data were analysed using the R statistical software R version 4.4.0. Participant characteristics were described using means and standard deviations (SD) for continuous data, and frequencies and proportions for categorical data. The variables included in the multivariable regression analysis were selected based on their association with contraceptive uptake from pre-existing literature (5,19–21).

Crude odds ratios (COR) with 95% confidence intervals (CI) and *p*-values were obtained after each predictor was assessed for its association with contraceptive uptake using bivariable logistic regression. The adjusted odds ratios (AOR) with 95% CI, and *p*-values were obtained from the multivariable logistic regression model, with statistical significance set at *p*-value < 0.05.

Collinearity among the predictors was tested using the variance inflation factor (VIF). A VIF of 10 or more was deemed to denote significant multicollinearity.

## 3. Results

Of the 1,274 caesarean deliveries performed at the study hospitals, 39 were excluded for data entry errors. These included one record with an age of 117 years, and 38 records in which women were recorded as having received both an intra-operative tubal ligation and another contraceptive method at one-year follow-up. A total of 1,044 women had complete data and were included in the complete-case analysis (Fig 1).

The mean participant age was 25.9 years (SD 6.3 years), with most mothers attending more than two antenatal care visits for their index pregnancy (Table 1). Half of all caesarean deliveries occurred at a regional or tertiary facility. Only 6.3% of women were offered a contraceptive method during their admission.

**Table 1.** Participant demographic table of women who underwent a caesarean delivery.

| CHARACTERISTIC | N=1,044<sup>1</sup> |
| --- | --- |
| **Age Group (Years)** |  |
| Mean age group (SD) | 25.9 (6.3) |
| 13-19 | 204 (19.5%) |
| 20-34 | 712 (68.2%) |
| 35+ | 128 (12.3%) |
| **Parity** |  |
| Mean Parity (SD) | 1.7 (2.0) |
| Nullipara | 363 (34.7%) |
| Multipara | 580 (55.6%) |
| Grand multipara | 101 (9.7%) |
| **Antenatal Care visits** |  |
| >2 visits | 942 (90.0%) |
| **Caesarean Delivery Type** |  |
| Planned | 156 (15.0%) |
| Emergency | 888 (85.0%) |
| **Caesarean Delivery Indication** |  |
| Antepartum haemorrhage | 117 (12.3%) |
| Prolonged labour | 591 (62.0%) |
| Uterine rupture | 29 (3.0%) |
| Fetal Indication | 75 (7.9 %) |
| Previous Caesarean delivery | 141 (14.8 %) |
| Missing | 91 |
| **Perinatal death** | 175 (17%) |
| **Post-op complications** | 107 (10.3%) |
| Missing | 45 |
| **Contraception Offered During Admission** | 60 (6.3%) |
| Missing | 94 |
| **Education Level** |  |
| None | 363 (35.3%) |
| Primary | 59 (5.7%) |
| Secondary | 381 (37.1%) |
| Tertiary | 225 (21.9%) |
| Missing | 16 |
| **Wealth Quintile** |  |
| Poorest | 43 (4.2%) |
| Poor | 49 (4.8%) |
| Middle | 99 (9.7%) |
| Rich | 231(22.7%) |
| Richest | 594 (58.5%) |
| Missing | 28 |
| **Marital Status** |  |
| Never married | 213 (20.7%) |
| Married | 814 (79.1%) |
| Widowed | 2 (0.2%) |
| Missing | 15 |
| **Facility** |  |
| District | 350 (34.0%) |
| Regional/ tertiary | 521 (50.0%) |
| Private/ non-profit | 173 (17.0%) |
| **Travel Time To Facility >2hrs** | 183 (18.0%) |
| Missing | 7 |
| <sup>1</sup>n (%) |  |

The contraceptive uptake was 48.5% in year five. The use of implants nearly doubled over this period (from 24.7% to 45.6%, *p*<0.001) while IUCD use declined from 40.3% in year one to 0.8% in year five (*p*<0.001) (Fig 2). There was no data for DMPA uptake in year one from the original cohort.

**Fig 2.**
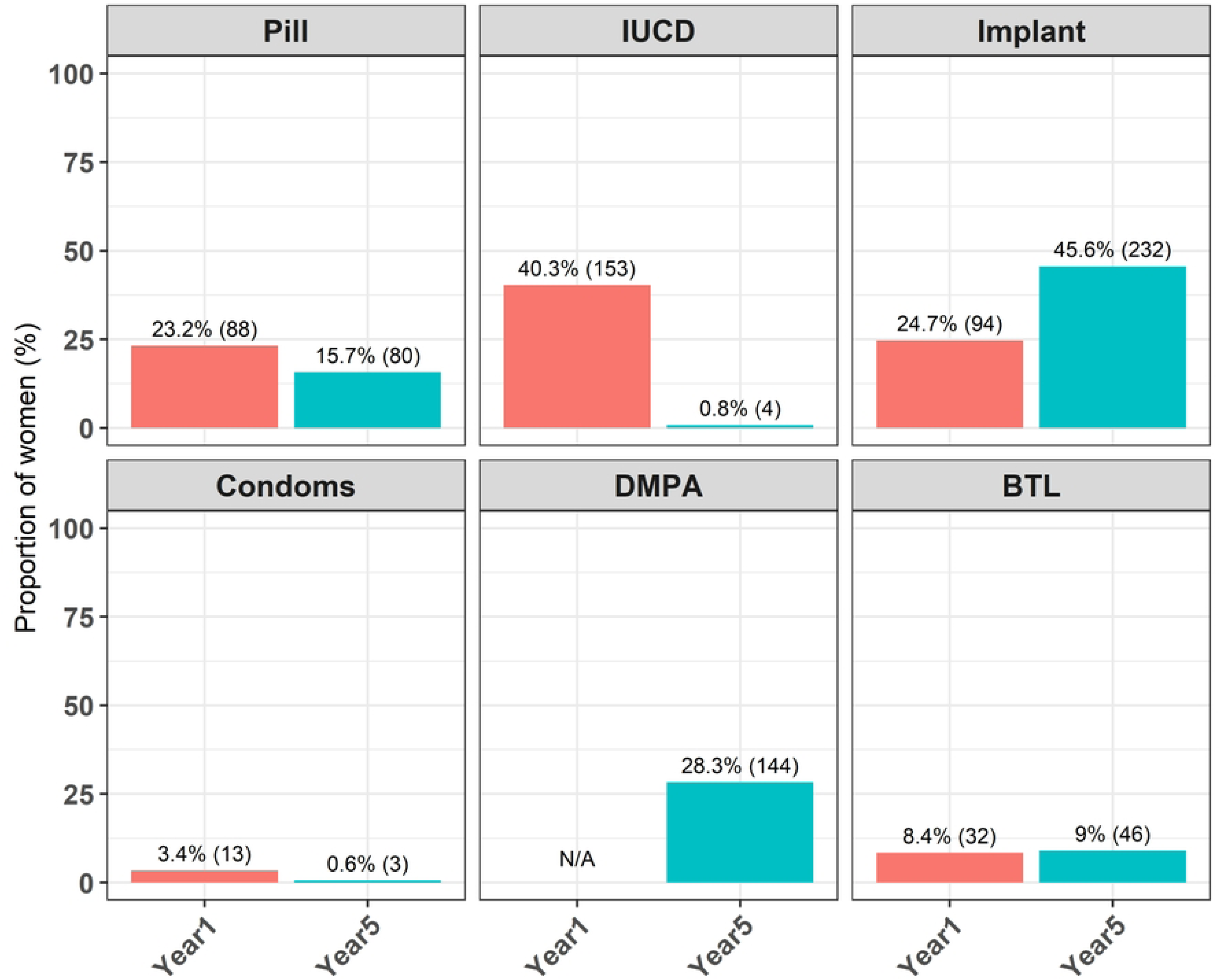
Proportion of women using modern contraceptives at one- and five-years post-caesarean delivery in Sierra Leone.

Delivery at a tertiary or regional facility was associated with 47% lower odds in contraceptive uptake compared to other facility types [aOR 0.53; 95% CI (0.34, 0.83)] (Table 2). However, women who attended more than two antenatal visits were nearly twice as likely to use a contraceptive method [aOR 1.96; 95% CI (1.19, 3.23)]. Additionally, offering a contraceptive method before discharge was significantly associated with a 2.44-fold increase in the uptake of a contraceptive method [aOR 2.44; 95% CI (1.05, 6.40)]. Although an increase in age was associated with a decreased uptake of contraception [aOR 0.98; 95% CI (0.95, 1.01)] and being rich was associated with a nearly two-fold increase in uptake [aOR 2.04; 95% CI (0.75, 5.47)], neither was statistically significant.

**Table 2.** Predictors associated with the uptake of modern contraceptive methods among women post-caesarean delivery.

| Variable | Category | Uptake of modern contraceptive methods |  |  |  |
| --- | --- | --- | --- | --- | --- |
|  |  | cOR <sup>a</sup> (95% CI <sup>a</sup>) | *P*-value | aOR <sup>a</sup> (95% CI) | *P*-value |
| **Age** | - | 0.97 (0.95, 0.99) | 0.002** | 0.98 (0.95, 1.01) | 0.199 |
| **Parity** | - | 1.00 (0.94, 1.07) | 0.974 | 1.09 (0.96, 1.24) | 0.183 |
| **Perinatal death** |  |  | 0.260 |  | 0.603 |
|  | None | Ref |  |  |  |
|  | Occurred | 0.81 (0.58-1.14) |  | 0.89 (0.57, 1.39) |  |
| **Education level** |  |  | 0.258 |  | 0.128 |
|  | None | Ref |  | Ref |  |
|  | Primary | 1.32 (0.74, 2.37) |  | 1.50 (0.72, 3.29) |  |
|  | Secondary | 1.35 (0.97, 1.78) |  | 1.47 (0.97, 2.24) |  |
|  | Tertiary | 1.16 (0.82, 1.64) |  | 1.64 (0.98, 2.77) |  |
| **Marital status** |  |  | 0.087 |  | 0.182 |
|  | Not married | Ref |  | Ref |  |
|  | Married | 0.72<br>(0.52,1.00) |  | 0.69 (0.45,<br>1.06) |  |
|  | Widowed | 327,278 (0.00,<br>NA) |  | 476,712<br>(0.00, NA) |  |
| **Wealth Quintile** |  |  | 0.015* |  | 0.677 |
|  | Poorest | Ref |  | Ref |  |
|  | Poor | 0.94 (0.40-2.18) |  | 1.94 (0.58,<br>6.61) |  |
|  | Middle | 1.50 (0.70-3.20) |  | 1.56 (0.53,<br>4.55) |  |
|  | Rich | 1.39 (0.71-2.75) |  | 2.04 (0.75,<br>5.47) |  |
|  | Richest | 0.96 (0.51-1.82) |  | 1.87 (0.66,<br>5.22) |  |
| **Contraceptive offered during admission** |  |  | 0.019* |  | 0.037* |
|  | Not offered | Ref |  | Ref |  |
|  | Offered | 2.22 (1.16,<br>4.25) |  | 2.44 (1.05,<br>6.40) |  |
| **Facility** |  |  | 0.042* |  | 0.018* |
|  | District | Ref |  | Ref |  |
|  | Regional/<br>Tertiary | 0.73 (0.55,<br>0.97) |  | 0.53 (0.34,<br>0.83) |  |
|  | Private/<br>non-profit | 1.03 (0.70,<br>1.54) |  | 1.00 (0.41,<br>2.64) |  |
| **>2 antenatal**<br>**visits** |  |  | 0.005** |  | 0.009** |
|  | ≤2 visits | Ref |  | Ref |  |
|  | >2 visits | 1.84 (1.22,<br>2.78) |  | 1.96 (1.19,<br>3.23) |  |
|  | <sup>a</sup> cOR = Crude Odds Ratio, aOR= adjusted Odds Ratio,<br>CI = Confidence Interval |  |  |  |  |

## 4. Discussion

This study examined the uptake of modern contraceptive methods and factors influencing their use within five years of caesarean delivery in Sierra Leone. Overall contraceptive uptake at five years was 48.5%, which is higher than the national contraceptive prevalence rate (CPR) of 29% (18). Unlike the national CPR, which reflects contraceptive use among all women of reproductive age (15–49 years) (22), our study only focused on women post-caesarean delivery, regardless of age. This difference may explain the higher uptake observed in our cohort. The increase over time may also partly reflect the 2016–2018 campaign by Marie Stopes Sierra

Leone (MSSL), a non-governmental organisation that has been providing quality and affordable sexual and reproductive health services in Sierra Leone since 1986 (23). The campaign, which included audio-based education and community engagement, was associated with a 40% increase in adult family planning clinic attendance and a doubling of adolescent clinic visits during promotion months.

Several factors known to affect contraceptive uptake include level of education, parity, exposure to audio information, and facility contact (17,20,24). Improvements in Sierra Leone’s female literacy, which increased from 44% in 2013 to 54% in 2019 (25), may have also contributed to the relatively high contraceptive uptake observed at five years, as more educated women tend to readily understand the benefits of contraceptive use (20,26). However, in contrast to prior studies, our study did not find a significant association between education level and contraceptive use.

Direct comparison of the overall contraceptive uptake between years one and five was limited by the absence of data on DMPA use at year one in the original cohort study. Excluding DMPA from the comparison could have resulted in an overestimation of the changes in uptake between the two years.

A notable finding from our study was that women who attended two or more ANC visits were significantly more likely to use modern contraceptives. This is consistent with findings from Uganda, where counselling during antenatal care was associated with a nine-fold increase in contraceptive adoption (27). Increased ANC attendance provides more opportunities for health education, including family planning counselling, which can empower women to make informed reproductive choices. This is particularly important in Sierra Leone, where 58% of women had not discussed family planning with either a fieldworker or a health provider in the year preceding the 2019 Sierra Leone Demographic Health Survey (SLDHS) (5).

Conversely, women who delivered at tertiary or regional referral hospitals were 45% less likely to use contraceptives. This could reflect limited provider–patient interaction during hospital stay, usually resulting from a human resource shortage. In Sierra Leone, tertiary facilities receive the highest referral rates (28), contributing to the increased workload on health providers. Although tertiary hospitals typically employ more skilled staff (29,30), high patient volumes, particularly in urban centres with accessible transportation, can reduce the time available for personalised care and counselling (28). A 2017 study done at Princess Christian Maternity Hospital reported that nearly half of the postpartum women reported that procedures were not explained to them, and 57% believed that staffing was inadequate (30). These findings highlight the need to strengthen patient engagement and counselling, especially in higher-level health facilities. The reduced uptake in tertiary settings could also be attributed to the MSSL campaign being primarily focused on the primary and community levels of care (23).

Regarding method-specific uptake, the IUCD was the most utilised method in the first year postpartum, likely due to the ease of intraoperative insertion. Another possibility would be the increased knowledge of IUCD. The 2023 Sierra Leone Obstetric Postpartum Programme showed the IUCD had the highest method information index (MII) of 89% (18), an indicator of counselling quality. However, by year five, IUCD use dropped to 0.8%, similar to the national use of IUCD of 0.4% (5). The reduction may reflect discontinuation due to side effects, which are the most common reason for contraceptive discontinuation in Sierra Leone (5). This was similarly reported in Uganda, where the fear of side effects was a barrier to the use of IUCDs (27).

By year five, implants were the most used method, surpassing DMPA uptake, which remains the most widely used contraceptive nationally and across Sub-Saharan Africa (5,31).

Although not statistically significant, women who were offered a contraceptive method before discharge were nearly 2.5 times more likely to adopt a method. This mirrors findings in Ethiopia where postpartum women who received counselling before discharge were more likely to use modern contraceptives at six weeks and six months postpartum (32). This underscores the importance of providing counselling before discharge, particularly where follow-up visits may be compromised by barriers such as transport costs (24,27). Notably, 17% of mothers in Sierra Leone do not receive a 48-hour postnatal check (5), further highlighting the need to strengthen counselling and service delivery before women leave the facility.

The study had notable strengths, including the relatively large sample size provided sufficient power for precise estimates. The inclusion of multiple hospitals from all regions of the country enhanced the generalizability of the findings. The long follow-up period, combined with low attrition rates, allowed for a reliable assessment of contraceptive uptake trends over time.

Furthermore, the analysis incorporated key covariates known to influence contraceptive uptake. Several limitations should be acknowledged. First, the use of secondary data restricted the analysis to variables collected in the original study, potentially excluding other important determinants of contraceptive use. Second, the presence of missing data may have introduced selection bias. In addition, the lack of data on DMPA use in year one may have led to an underestimation of contraceptive uptake. Finally, the absence of data from intermediate time points limited our ability to perform a detailed trend analysis over the five-year period.

## 5. Conclusion

Family planning plays a critical role in reducing maternal morbidity and mortality. This study identified factors associated with contraceptive uptake following caesarean delivery, with frequent antenatal care attendance and the type of health facility being significant predictors. These findings highlight the importance of strengthening provider–patient interactions to support informed contraceptive choices. Future research should incorporate qualitative methods to explore the reasons behind discontinuation and deepen understanding of contextual and individual-level barriers to sustained contraceptive use.

## Data Availability

All relevant data underlying the findings of this study are available in the Zenodo repository at https://doi.org/10.5281/zenodo.18871119

## Acknowledgements

We are grateful to the study participants for their willingness to take part and to the data collection team for their dedication and effort in collecting data. We would also like to thank the directors of the nine health facilities that allowed us to conduct the study in their institutions.

## Notes

### Competing Interest Statement

The authors have declared no competing interest.

### Funding Statement

Yes

### Author Declarations

The original study was conducted in accordance with the Declaration of Helsinki and approved by the Sierra Leone Ethics Committee and the Regional Committees for Medical and Health Research Ethics in Central Norway (ethical clearance number 2016/1163)

## References

1. WHO, UNICEF, UNFPA, World Bank Group, UNDESA/Population Division. Trends in maternal mortality 2000 to 2023 [Internet]. 2025 Apr [cited 2025 Dec 29]. Available from: https://data.unicef.org/resources/trends-in-maternal-mortality-2000-to-2023/

2. United Nations Children’s Fund. Maternal mortality rates and statistics - UNICEF DATA [Internet]. 2025 [cited 2025 Oct 9]. Available from: https://data.unicef.org/topic/maternal-health/maternal-mortality/

3. Geneva: World Health Organization. Trends in maternal mortality 2000 to 2020: Estimates by WHO, UNICEF, UNFPA, World Bank Group and UNDESA/Population Division [Internet]. 2023 [cited 2025 Jul 9]. Available from: www.who.int/publications/i/item/9789240068759

4. World Health Organization. Maternal mortality [Internet]. 2025 [cited 2025 Jul 9]. Available from: https://www.who.int/news-room/fact-sheets/detail/maternal-mortality

5. Statistics Sierra Leone, ICF. Sierra Leone Demographic and Health Survey 2019. Freetown; 2020 Oct.

6. World Health Organization. World Health Organization. 2024 [cited 2025 Jul 9]. Adolescent pregnancy. Available from: https://www.who.int/news-room/fact-sheets/detail/adolescent-pregnancy#:∼:text=Key%20facts,births%20(1%2C2)

7. Shafiq Y, Caviglia M, Juheh Bah Z, Tognon F, Orsi M, Kamara AK, et al. Causes of maternal deaths in Sierra Leone from 2016 to 2019: Analysis of districts’ maternal death surveillance and response data. BMJ Open. 2024 Jan 12;14(1).

8. Logstein E, Torp R, Ashley T, Kamara MM, Koroma AP, Dumbuya AB, et al. Long-term maternal outcomes 5 years after cesarean section in Sierra Leone: A prospective cohort study. International Journal of Gynecology and Obstetrics. 2025 Mar 1;168(3):1210–20.

9. Cunningham F, Leveno k. Williams Obstetrics 25th Ed. 25th ed. Cunningham F, editor. McGraw-Hill Education; 2018. 989–1025 p.

10. Bauserman M, Nowak K, Nolen TL, Patterson J, Lokangaka A, Tshefu A, et al. The relationship between birth intervals and adverse maternal and neonatal outcomes in six low and lower-middle income countries. Reprod Health. 2020 Nov 1;17.

11. Garg B, Darney B, Pilliod RA, Caughey AB. Long and short interpregnancy intervals increase severe maternal morbidity. In: American Journal of Obstetrics and Gynecology. Mosby Inc.; 2021. p. 331.e1–331.e8.

12. van Duinen AJ, Westendorp J, Kamara MM, Forna F, Hagander L, Rijken MJ, et al. Perinatal outcomes of cesarean deliveries in Sierra Leone: A prospective multicenter observational study. International Journal of Gynecology and Obstetrics. 2020 Aug 1;150(2):213–21.

13. Chola L, McGee S, Tugendhaft A, Buchmann E, Hofman K. Scaling up family planning to reduce maternal and child mortality: The potential costs and benefits of modern contraceptive use in South Africa. PLoS One. 2015 Jun 15;10(6):1–13.

14. Ezzati Majid, Lopez A, Rodgers A, Murray C. Comparative quantification of health risks : global and regional burden of disease attributable to selected major risk factors. Vol. 1. World Health Organization; 2004.

15. Donnelly J. How did Sierra Leone provide free health care? The Lancet. 2011;377(9775):1393–6.

16. Government of Sierra Leone. FP2030 Commitment. 2023.

17. Sserwanja Q, Nuwabaine L, Kamara K, Musaba MW. Prevalence and factors associated with utilisation of postnatal care in Sierra Leone: a 2019 national survey. BMC Public Health. 2022 Dec 1;22(1).

18. Family planning 2030. Exploring Opportunities for mCPR Growth in Sierra Leone [Internet]. 2024 Apr [cited 2025 Jul 9]. Available from: https://www.fp2030.org/app/uploads/2023/08/Sierra-Leone-FP-Opportunity-Brief.pdf

19. Forty J, Rakgoasi SD, Keetile M. Patterns and determinants of modern contraceptive use and intention to usecontraceptives among Malawian women of reproductive ages (15–49 years). Contracept Reprod Med. 2021 Dec;6(1).

20. Osborne A, Bangura C. Proximal factors influencing the likelihood of married and cohabiting women in Sierra Leone to use contraceptives. A cross-sectional study. Contracept Reprod Med. 2024 Dec 1;9(1).

21. Sserwanja Q, Nuwabaine L, Kamara K, Musaba MW. Determinants of quality contraceptive counselling information among young women in Sierra Leone: insights from the 2019 Sierra Leone demographic health survey. BMC Womens Health. 2023 Dec 1;23(1).

22. The world bank. The World Bank in Malawi . 2020.

23. Marie Stopes International. Marie Stopes International. 2018 [cited 2025 Jul 9]. Reframing the conversation and removing barriers: what makes a promotional campaign on contraception a success with urban youth. Available from: https://www.msichoices.org/wp-content/uploads/2023/10/brief_9_promotional-campaign.pdf

24. Sylvain MH, Valens R. Factors associated with postpartum family planning use in Rwanda. Contracept Reprod Med. 2024 Dec 1;9(1):1.

25. National Statistical Office. Malawi Demographic and Health Survey 2015-16. National Statistics Office The DHS Program [Internet]. 2015;1–658. Available from: http://dhsprogram.com/pubs/pdf/FR319/FR319.pdf

26. Olarenwaju Saheed J, Vaughan T, Adeniyi M, Ahmed A, Sule-Odu L, Badmaasi-Abdulraheem T, et al. Patterns And Predictors Of Contraceptive Uptake Among Women Attending Family Planning Clinic In A Tertiary Health Facility In South-West Nigeria: A 10-Year Review. African Journal of Research in Medical and Health Sciences. 2024;2(1):12–9.

27. Nakiwunga N, Kakaire O, Ndikuno CK, Nakalega R, Mukiza N, Atuhairwe S. Contraceptive uptake and associated factors among women in the immediate postpartum period at Kawempe Hospital. BMC Womens Health. 2022 Dec 1;22(1).

28. Youkee D, Bangura T, O’Neill K, Hartshorn L, Samura S. An Observational Study of a National Referral System: An Analysis of 14,266 Referrals in Sierra Leone [Internet]. 2020. p. 1. Available from: https://www.researchsquare.com/article/rs-52747/v1

29. Willie MJ, George AM, George DR. The Imperative for Expanded Specialized Nursing Personnel in Sierra Leone: A Case Study of Four Referral Hospitals. International Journal of Research Publication and Reviews Journal homepage: www.ijrpr.com [Internet]. 2024 Dec;5:561–71. Available from: www.ijrpr.com

30. Mclellan A, Van Ham PT, Sidney D, Aden A, Lacroix A, Edem-Hotah J. Examining person-centred maternal care services at the Princess Christian Maternity Hospital, Freetown, Sierra Leone. Afr J Midwifery Womens Health. 2022 Apr 2;16(2):1–12.

31. World Health Organization. Sierra Leone Contraception within the context of adolescents’ sexual and reproductive lives: Country profile SOCIO-DEMOGRAPHIC CHARACTERISTICS [Internet]. 2020. Available from: http://data.uis.unesco.org

32. Mruts KB, Tessema GA, Gebremedhin AT, Scott JA, Pereira G. The role of family planning counselling during maternal and child health services in postpartum modern contraceptive uptake in Ethiopia: A national longitudinal study. PLOS Global Public Health. 2022 Aug 3;2(8).

33. National statistical office (NSO) [Malawi], ICF. Malawi Demographic Health Survey 2015-16. Zomba, Malawi and Rockville Maryland, USA; 2017.

